# Using virtual reality to investigate bias and discrimination in clinical decision making: A proof of concept study

**DOI:** 10.1101/2025.09.26.25336643

**Authors:** Zoe Chui, Helen Welsh, Rebecca Rhead, Lucia Valmaggia, Juliana Onwumere, Lucy Ensum, Stephani Hatch

## Abstract

**Background:** Biases held by healthcare practitioners can shape clinical interactions, leading to discrimination and poor patient outcomes. Traditional methods, such as vignettes or self-report measures, often lack ecological validity.

**Objective:** We evaluated a novel VR simulation designed to explore bias and discrimination in clinical decision-making.

**Methods:** Thirty-five healthcare practitioners across 14 NHS trusts completed an in-person VR simulation and provided feedback on realism, immersion, and usability via surveys and open-ended responses.Patients in the simulation had intersecting sociodemographic characteristics (race, gender, migration status).

**Results:** Most participants rated the VR environment as at least moderately realistic (88.6%), with 42.9% rating it very or extremely realistic. Nearly half (45.7%) reported consistency with real-world experience. Open-text feedback highlighted strengths in realism, immersion, and educational potential, with suggestions to improve dialogue flexibility.

**Conclusions:** VR is a feasible, immersive, and scalable tool for investigating bias in clinical decision-making.

**Graphical Highlights:**

- Virtual reality (VR) is feasible for studying bias in clinical decision-making.
- VR simulations capture intersecting patient characteristics, including race, gender, and migration status.
- Participants rated the VR environment as realistic, immersive, and usable.
- VR provides a scalable, controlled platform for research and professional training on bias and discrimination.

## Introduction

### Background

In healthcare settings, biased attitudes and beliefs held by practitioners towards different groups of patients can shape clinical interactions, outcomes, and levels of service satisfaction. This can also lead to direct and indirect acts of discrimination. In England and Wales, discrimination is defined as treating a person unfairly because of characteristics protected by the Equality Act 2010, which, in a healthcare context, can result in poor care outcomes for patients [1–3].

Research on identifying and reducing clinician bias and discrimination in healthcare has largely relied on self-report questionnaires and clinical vignettes [3, 4]. These methods lack ecological validity (i.e., the degree to which a study’s methods reflect the complexity of real world situations) and are unable to capture the full range of clinician–patient interactions that occur in practice. While some studies have attempted to address these limitations using video-based cases and role-play scenarios, these approaches remain limited in its ability to standardise patient behaviour across all aspects of encounters [5, 6].

Traditional approaches to clinical education and bias training through simulated patients, role-play, and 2D video case studies also present limitations. Live simulation provides high fidelity but is resource-intensive and costly to scale [7,8], whereas 2D video lacks immersion and interactivity. Virtual reality (VR) offers a middle ground: studies suggest it can achieve comparable learning outcomes to high-fidelity simulation but at lower cost, while enhancing engagement and immersion compared with video-based learning [9 -12].

Bias and discrimination have typically been investigated using written vignettes, standardised patients, and role-play exercises [13,14]. While valuable, these methods often lack scalability and ecological validity. Emerging applications of VR in health professions education have demonstrated promise for enhancing realism and directly influencing bias related attitudes and behaviours. For example, embodiment in avatars of different racial backgrounds has been shown to reduce implicit racial bias [15, 16, 17]. This underscores the potential of VR as a novel platform for both investigating and addressing bias in clinical decision-making.

VR may be particularly advantageous for studying socially undesirable or harmful behaviours, such as discrimination, since well-designed simulations can evoke responses that closely mirror real world behaviour [18]. VR can also be used to examine how intersecting sociodemographic characteristics (e.g., gender, race, ethnicity, and class) shape clinical interactions. These intersecting characteristics influence not only how clinicians perceive and interact with patients, but also the treatment recommendations they make [19, 20].

Despite these benefits, VR remains underutilised in non-experimental social science studies of bias and discrimination. Given the potential of VR to generate ecologically valid evidence on this pressing source of health inequality, it is important to establish the feasibility of this method for researching bias and discrimination in clinical decision-making.

### Aims

The Tackling Inequalities and Discrimination Experiences in Health Services (TIDES) study developed a VR simulation to address limitations of existing methods for studying bias and discrimination. The overarching aim of TIDES is to understand how witnessed and experienced discrimination contributes to health inequalities. This study specifically aimed to evaluate the feasibility of VR in simulating clinical scenarios and gathering participant feedback on realism, usability, and its potential for investigating bias.

## Methods

### Study Setting

The study was conducted at King’s College London by the TIDES study team (Department of Psychological Medicine) and the VR Research Lab (Department of Psychology). A trained researcher facilitated VR sessions, supported by a VR lab developer who managed software updates and exported headset data.

### Participants

The VR study built on the 2019 TIDES survey of harassment and discrimination among staff in London NHS trusts [21]. The survey included 931 healthcare practitioners. Purposive sampling ensured diversity across occupation, gender, ethnicity, and migration status. Participants consenting to be recontacted were invited to join the VR study. Exclusion criteria included seizure or vestibular disorders or uncorrected hearing or vision issues [22, 23].

### VR Equipment

The simulation, built by Dan Archer of Empathetic Media using Unity3D, ran on Oculus Quest headsets. Participants interacted using a hand-held controller.

### VR Simulation Development

The VR simulation was developed iteratively with the TIDES team, clinicians, and VR specialists, following guidelines for immersive VR in cognitive neuroscience [24]. Realism was a central goal: the simulation recreated a primary care consultation room from a first-person perspective, with interactive elements such as virtual hands and a keyboard (Figure 1). Dialogue trees co-developed with clinicians comprised 32 questions across eight categories.

**Figure 1.**
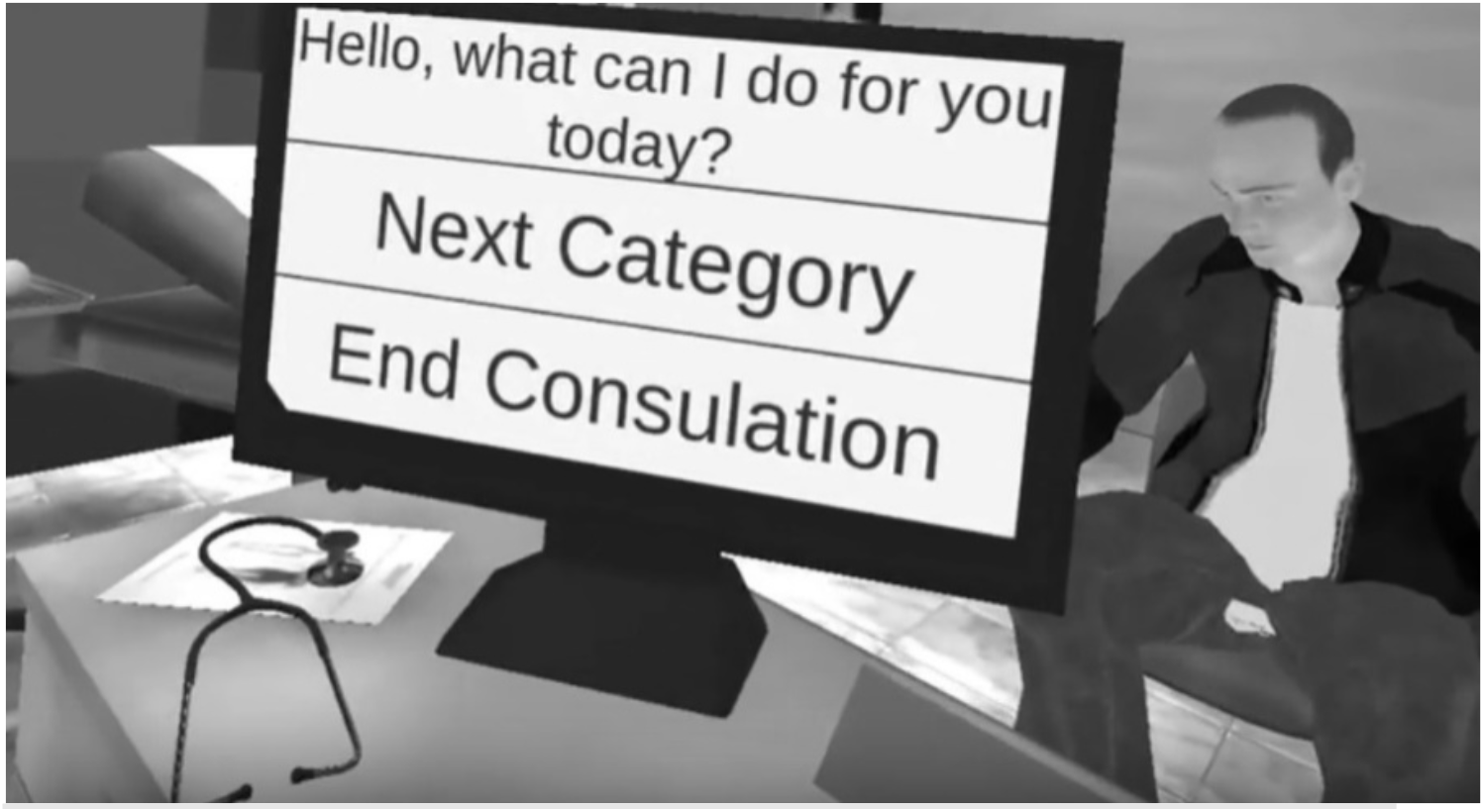
An example of the participant’s view within the VR simulation (Black and white in print)

Patient avatars reflected intersecting sociodemographic characteristics, including gender, race, and migration status (Figure 2). Intersectionality was considered so participant decisions could be observed relative to overlapping social identities. The design balanced technical fidelity (graphics, responsiveness) with social and cognitive realism (interaction with avatars, clinical decision-making) [25, 26]. Participant feedback guided refinements in conversational flow, empathy, and usability.

**Figure 2.**
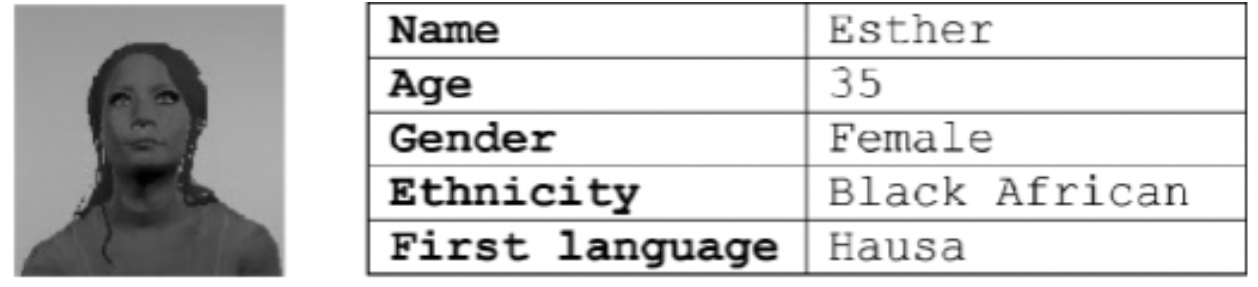
An example of a *virtual patient profile (Black and white in print)*

### Procedure

Participants were first provided with a digital information sheet and could ask questions via email before participation. Those interested attended a King’s College London campus of their choice, where they received a paper copy of the information sheet and consent form. The researcher reviewed these documents with each participant, answered questions, and emphasised that data would be stored securely at King’s College London and accessed only by the TIDES team. Participants who agreed signed a digital version of the consent form.

They then completed a short sociodemographic survey on a tablet, covering age, education, household composition, job title, NHS Trust, occupational group, pay band, income, years in healthcare, patient contact, and NHS Equality and Diversity training (Supplementary Table 1). Participants received a brief description of each virtual patient (Figure 2) and estimated expected consultation length. Meanwhile, the researcher sanitised and prepared the Oculus Quest headset and controller. Because the headset obscured physical surroundings, care was taken to ensure a safe environment free of obstacles. All participants remained seated throughout the simulation.

After adjusting the headset for comfort, participants completed a short tutorial to learn the controller and navigation. The main simulation lasted 30–45 minutes. Each virtual consultation was capped at ten minutes, displayed on a virtual wall clock (Figure 1). Participants assessed each patient and recorded treatment recommendations using a virtual dictaphone.

Following each consultation, participants completed a short in-VR survey rating the patient’s personality, probable behaviour, social roles, and overall health. After the simulation, the researcher assisted participants in removing the headset, which was sanitised again.

Participants remained seated briefly to readjust and were offered refreshments to mitigate potential motion sickness [28]. Finally, they completed a feedback survey on a tablet, taking approximately five minutes.

### Measures

The feedback survey consisted of the following three questions which participants responded to using a five-point Likert scale:

1. “*How real did the virtual world seem to you*?” (0 = Not real at all to; 4 = Extremely real)
2. “*How much did your experience in the virtual environment seem consistent with your real-world experience*?” (0 = Not consistent; 4 = Extremely consistent)
3. “*How aware were you of the real-world surroundings while navigating the virtual world? (i*.*e. sound, room temperature, other people, etc*.)?” (0 = Not aware at all; 4 = Extremely aware)

### Analysis

Descriptive statistics were calculated to describe participants’ responses to the three quantitative items in the feedback survey. Analyses were conducted in Stata 17 [29].

### Ethical Approval

Ethical approval was granted by the King’s College London Research Ethics Committee for Psychiatry, Nursing and Midwifery (HR-17/18-4629) and NHS Health Research Authority (18/HRA/0368).

## Results

### Sample Characteristics

A total of 35 healthcare practitioners across 14 NHS trusts participated. Twenty participants were qualified nurses, seven were medical doctors, and the remaining were allied health professionals, including therapists, psychologists, and pharmacists. Participants’ salary bands ranged from band 5 (newly qualified staff) to band 8 (managers with over five years’ experience). Twenty-seven participants were female, and the remainder were male. Ethnic backgrounds included White British or Irish (14 participants), Black African or Asian ethnic groups (12) and other White ethnic groups (9 participants). The mean age of participants was 35.3 years.

### Realism, consistency with reality and immersion

As shown in Figure 3, most participants (88.6%) rated the VR simulation as at least moderately real, with 42.9% describing it as very or extremely real. Nearly half (45.7%) reported that the simulation was very or extremely consistent with their real-world experience, while 51.4% reported being only slightly or not at all aware of their real-world surroundings during the task, suggesting a high degree of immersion.

**Figure 3.**
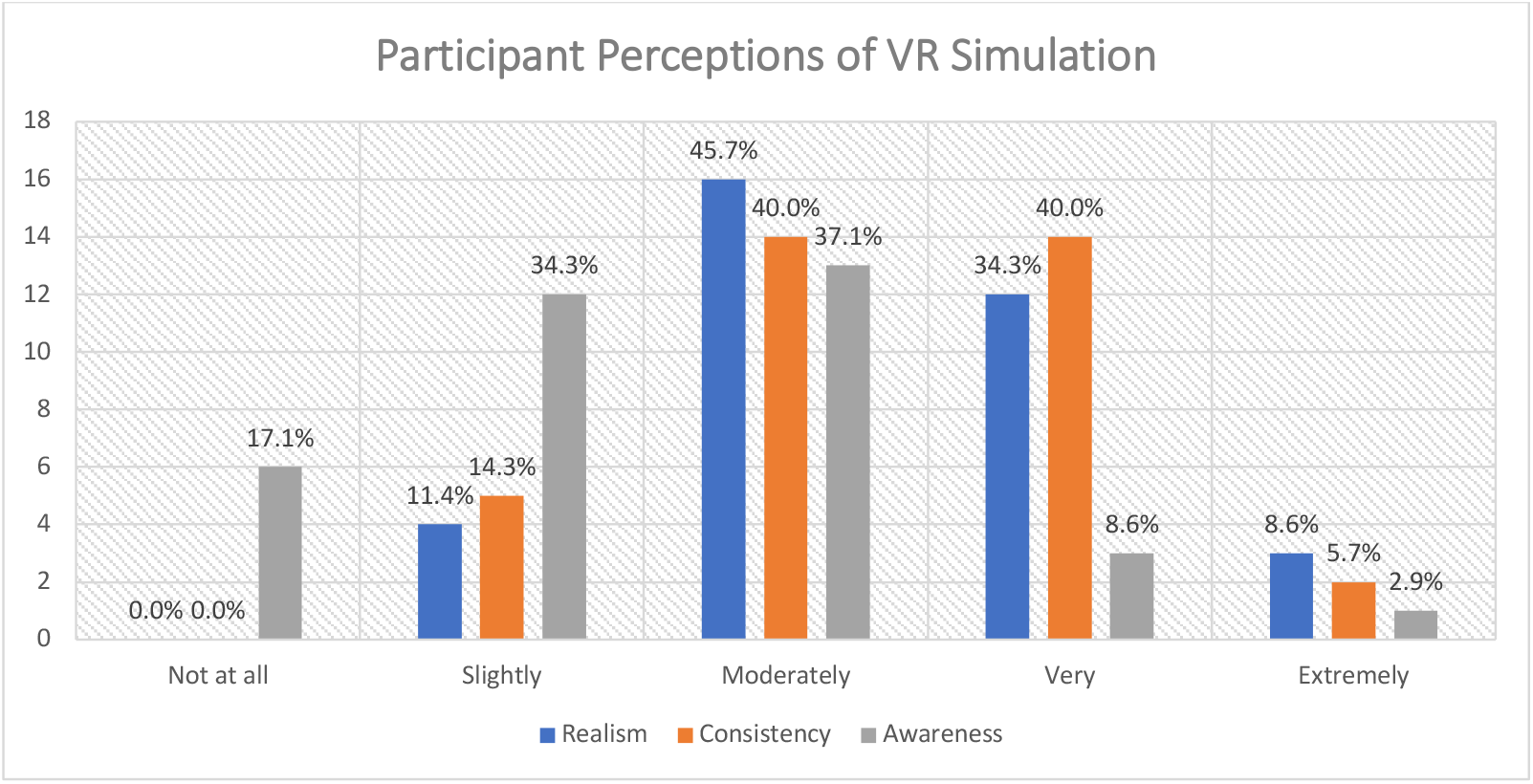
Participants’ perceptions of the realism of the VR simulation, consistency between the VR simulation and their real-world experience, and awareness of real-world surroundings while immersed in the VR simulation. (Color online)

### Open-ended feedback on technical aspects

Participants’ qualitative feedback emphasised usability and technical feasibility (Textbox 1). No participants reported motion sickness or physical discomfort. Many described the simulation as realistic, and most found the overall length appropriate for a training exercise. Some reported initial difficulty using the virtual dictaphone to record treatment recommendations, but these issues were quickly resolved with researcher assistance. This highlights the importance of brief, structured tutorials for maintaining usability in VR based studies.

### Open-ended feedback on content

Feedback on the VR simulation content was broadly positive, with participants noting that consultation questions were clear, comprehensive, and clinically appropriate (Textbox). Nonetheless, several participants identified areas for improvement. Some reported that fixed, pre-determined questions felt restrictive, limiting opportunities for follow-up or elaboration and constraining more natural consultations. Others had difficulty navigating question sets, commenting that not all prompts were visible within each category, occasionally causing repetition and disrupting flow. A further challenge concerned treatment recommendations; participants found it difficult to decide without additional diagnostic information, such as test results. This was particularly evident among nurses in specialist roles, who felt independent recommendations were unrealistic compared with their usual practice.

Taken together, these points suggest future iterations could benefit from more complex dialogue trees, improved navigation to display all options, and supplementary clinical data to support decisions. Matching participants to patients relevant to their professional expertise may also enhance ecological validity.

Analysis of open-ended responses further highlighted several themes. Usability was prominent, with most participants finding the system intuitive, though a minority required clarification on tasks like the virtual dictaphone. Realism and immersion were frequently mentioned; participants consistently described the environment and avatars as realistic, though greater conversational flexibility would strengthen authenticity. Another theme concerned empathy and communication, as fixed response options limited opportunities to demonstrate sensitivity and rapport. Finally, participants emphasised the simulation’s potential as a training tool for raising awareness of bias and discrimination in healthcare decision-making.

These findings reinforce the feasibility of VR for examining bias in clinical contexts while pointing to refinements needed to balance ecological and cognitive validity [30].

**Textbox 1**. A subset of direct quotes from open-ended feedback provided by participants

**Technical aspects of the VR simulation**

**Positive**

- Everything was clear, length perfect, good instructions provided by staff.
- Overall, I enjoyed the experience, felt realistic and would be very good for training purposes for doctors of all grades.
- Overall experience was very positive, very easy and very realistic.

**Negative**

- The only part that was slightly unclear to me personally was the use of the dictaphone for recommendations prior to the prompt by VR software.

**Content of the VR simulation**

**Positive**

- Questions were relevant based on the presentation of the patient, with the depressed client I think it captured a realistic tone of voice and body language one would see.
- I liked being led in the questions. I might naturally explore more around the questions in response to how the person answered, but understand that this is difficult to incorporate in the VR experience, but being able to talk out my train of thought and having freedom to give a treatment plan allowed space to verbalise this.
- The questioning was elaborate enough in terms of the presenting complaints.

**Negative**

- It would be helpful to see the questions in each category in advance so that you can plan the consultation in your head as it progresses. Some of the questions and patient answers were somewhat repetitive, and did not allow for empathetic responses/follow-up questions to what the patient had said.
- The questions seemed unintuitive and did not follow in a way that I would usually put them.
- It would be useful to be able to go back to a section. It would be useful to have examination findings e.g., back exam, PHQ9 score
- I think different medical backgrounds will likely have a different response to the patients. The only issue I had was making the clinical decisions at the end as this is something that I usually discuss with the doctor.

## Discussion

VR remains underutilised in research examining socially undesirable behaviours, such as bias and discrimination, within clinical decision-making. Compared to self-report questionnaires, vignettes, and role-play, VR enables the creation of controlled yet realistic scenarios where patient behaviour and the clinical environment can be standardised. Unlike role-play with actors, VR is less resource-intensive and more scalable, while offering greater immersion and ecological validity than 2D video-based approaches [12]. VR occupies a middle ground between low-cost but limited modalities and high-fidelity but costly live simulations [7–10].

The aim of this study was to evaluate healthcare practitioners’ feedback on a novel VR prototype developed to explore bias and discrimination in clinical encounters. Thirty-five participants rated the simulation as realistic and immersive, supporting its feasibility for investigating bias. While not designed to directly measure bias, the study demonstrates that VR can elicit meaningful reflections on realism, usability, and empathy, essential precursors for developing tools for bias research and training.

A key contribution of this work lies in the development process. The simulation was designed with attention to realism in patient presentation and decision-making prompts, as well as intersectionality in patient identities and contexts. These design choices are critical, as the effectiveness of VR depends not only on technical fidelity but also on capturing the social and interpersonal dynamics of practice [25]. By foregrounding equity and intersectionality, VR can extend beyond technical skills training into complex domains such as bias, discrimination, and healthcare inequalities [14,15,17].

The findings also point to VR’s potential as an educational tool. Diagnostic errors in primary and secondary care can have harmful consequences for patients, practitioners, and health services [31,32]. Evidence suggests ethnic minority patients face disproportionate risks of complications and adverse events [33]. VR exposes practitioners to diverse, intersectional patient scenarios safely, helping build awareness of bias and supporting equitable clinical decision-making. Previous research shows implicit bias affects clinical interactions [34,35], but traditional studies often assess single social identities. VR allows interaction with patients possessing multiple intersecting identities, offering new insights into health inequalities.

Despite its promise, the study highlights limitations. Fixed response options constrained dialogue and empathetic engagement, and participants noted challenges navigating questions and making treatment recommendations without additional diagnostic information. These concerns reflect broader design challenges in VR healthcare training: ensuring ecological validity while preserving cognitive validity [26]. Such limitations provide guidance for refinement, including more flexible dialogue trees, supplementary clinical data, and scope-sensitive case assignments.

## Strengths and limitations

This study demonstrated the feasibility of using VR to simulate complex clinical interactions while balancing methodological rigour with accessibility. By collecting both quantitative and qualitative data, we were able to evaluate immersion, realism, and user experience alongside practical considerations for clinical decision making. Although not within the scope of this paper, these data also provide opportunities for future analyses of how bias may influence treatment recommendations, as well as conversation analysis of clinician–patient interactions. A further strength was the accessibility of the prototype: the interactive features required minimal training, relying on natural speech and a simple point and click mechanism with the hand-held controller. This simplicity reduced participant burden, minimised the risk of motion sickness, and made the simulation suitable for diverse healthcare professionals.

Several limitations should also be noted. Participants were restricted to asking predetermined questions, since programming naturalistic responses to unrestricted input was not technically feasible within this prototype. This limitation reduced opportunities to observe potential indicators of bias in how clinicians select or phrase questions. Second, participants interacted with only three virtual patients to limit the risk of VR related discomfort with prolonged exposure, which constrained the range of scenarios available for analysis. Third, the rapid pace of VR hardware development posed challenges for implementation. Although the Oculus Quest was appropriate at the time of development, frequent software updates, including those associated with the transition from Facebook to Meta, created practical obstacles. Nonetheless, the simulation was designed with transferable software that remains compatible with newer headsets, supporting future adaptation and delivery.

## Further research

Whilst participants in this study rated the VR simulation as highly immersive, future research should build on these findings by expanding both the technical capabilities and the scope of VR simulations. Advances in hardware now allow for greater interactivity, such as manipulating objects or conducting physical examinations, which could enhance ecological validity. Simulations situated in diverse clinical contexts, for example inpatient wards, outpatient clinics, or community services, would enable exploration of how organisational and environmental factors shape decision making. In addition, future studies should examine decision making with patients varying across a broader range of protected characteristics, such as disability, age, or sexual orientation, and presenting with both physical and mental health conditions. Such work would extend the utility of VR for investigating bias and inform the design of training interventions that prepare clinicians to deliver equitable care across diverse patient populations.

## Conclusions

This proof of concept study demonstrates the feasibility and potential value of using VR to simulate complex clinical encounters for examining bias and discrimination in healthcare. Participants reported high levels of realism, immersion, and usability, indicating that VR can provide a controlled yet authentic platform for exploring clinician–patient interactions and eliciting meaningful feedback on decision making processes. By incorporating intersectional patient identities, the simulation also highlights how multiple social characteristics can influence clinical reasoning, offering a novel approach to studying bias that complements traditional methods such as vignettes, role-play, and 2D video.

The development process, grounded in collaboration between clinicians, psychologists, and VR specialists, ensured that both technical and social realism were central to the simulation. Feedback from participants further identified opportunities for refinement, such as greater flexibility in dialogue, enhanced access to clinical information, and better alignment of patient scenarios with practitioner expertise. These insights underscore how VR not only serves as a research tool but also has potential as an educational and training platform for healthcare professionals, supporting the development of skills in equitable decision making and empathy.

Future research can build on this foundation by expanding the scope and interactivity of VR simulations, diversifying clinical contexts, and including patients with a wider range of protected characteristics and health conditions. As VR technology becomes increasingly accessible and sophisticated, such applications have the potential to enhance evidence-based interventions, improve training in bias reduction, and ultimately contribute to more equitable healthcare delivery.

This study provides strong evidence that VR is a feasible and innovative method for investigating bias and discrimination in clinical decision making. By combining immersion, interactivity, and intersectional patient representation, VR offers a scalable, ecologically valid, and practical tool for both research and professional training in healthcare settings.

## Data Availability

Data is available for research purposes upon request. All requests are reviewed by the study data committee. To apply for access to this data please contact

## Acknowledgements

This study was conducted by the TIDES team members and colleagues at the VR Research Lab, King’s College London. We thank Luke Connor and Ruth Mintah for their assistance with developing the study protocol and draft manuscripts; Kate Rimes, Sarah Dorrington; and Catherine Polling for their clinical expertise during the script writing phase; Dan Archer from Empathetic Media for building the simulation; and Jerome Di Pietro for technical support during the recruitment phase.

---

**Supplementary Table 1.** Summary of sociodemographic survey items.

| Survey Item | Question | Response Options |
| --- | --- | --- |
| Date of Birth | “What is your date of birth?” | Free text |
| Age | “What was your age on your last birthday?” | Free text |
| Educational Attainment | “What is the highest level of education you have achieved?” | Below GCSE, GCSE or equivalent, A level or equivalent, HND, BTEC, Undergraduate (MBBS or MBBS BSc), Undergraduate (BSc/BA), Postgraduate (PG Cert), Postgraduate (MSc), Postgraduate (PhD) |
| Household | “In which of these ways do you occupy your current house?” | Own outright, Buying with the help of a mortgage or loan, Shared ownership (part-owned, part-rented), Rent (private), Rent (housing association or local authority), Keywork/student accommodation, Live with parent(s), Other (please specify) |
|  | “How many people are currently living in your household, including yourself?” | Free text |
|  | “Of these, how many are children?” | Free text |
|  | “Of these, how many are adults?” | Free text |
|  | “Of the adults, how many bring income into the household?” | Free text |
|  | “Which of these categories best describes your total combined household income for the last year?” | £0, £10k – 19k, £20k – 29k, £30k – 39k, £40k – 49k, £50k – 59k, £60k – 69k, £70 – 79k, £80k – 89k, £90k – 99k, £100k – 149,999, £150k or more, Prefer not to say |
| Job Title | “What is your full job title?” | Free text |
| NHS Trust | “Which NHS Trust is your main job/training based at?” | Barking, Havering & Redbridge University Hospital; Barnet, Enfield & Haringey Mental Health Trust; Barts Health; Camden & Islington Foundation Trust etc. |
| Occupational Group | “What is your occupational group?” | Allied health professional/Healthcare scientist or technician, Medical, Nursing, Healthcare assistant/Nursing assistant, Psychotherapy/Psychology, Ambulance (operational), Other (please specify) |
| Pay Band | “What is your Agenda for change pay rate?” | Band 1, 2, 3, 4, 5, 6, 7, 8, 9, N/A |
| Income | “How much did you earn from your primary/main employer before taxes and deductions last year?” | £0, £10k – 19k, £20k – 29k, £30k – 39k, £40k – 49k, £50k – 59k, £60k – 69k, £70 – 79k, £80k – 89k, £90k – 99k, £100k – 149,999, £150k or more, Prefer not to say |
|  | “Did you receive income last year from any other sources (e.g. HCA/locum)?” | Yes, No |
|  | “What was the source of income?” | Free text |
|  | “How much did you receive from this income last year?” | £0, £10k – 19k, £20k – 29k, £30k – 39k, £40k – 49k, £50k – 59k, £60k – 69k, £70 – 79k, £80k – 89k, £90k – 99k, £100k – 149,999, £150k or more, Prefer not to say |
| Years Worked in Healthcare | “How many years have you worked in health care?” | Free text |
| Patient Contact | “Do you have contact with patients?” | Yes, No |
| NHS Equality and Diversity Training | “Have you completed the NHS Equality and Diversity training?” | Yes, No |
|  | “How did you complete your training?” | Online, In person |
|  | “When did you last take part in this training?” | Less than 12 months ago, More than 12 months ago but less than 3 years, 3 or more years ago |

## Notes

**Funding** This paper represents independent research funded by the Wellcome Trust [203380/Z/16/Z] and the Economic and Social Research Council [ES/V009931/1]. J.O. is part-funded by the NIHR Biomedical Research Centre [BRC-1215–20018] at South London and Maudsley NHS Foundation Trust. S.L.H. and R.R. are supported by the Economic and Social Research Council Centre for Society and Mental Health at King’s College London [ES/S012567/1] and by UK Research and Innovation [MR/Y030788/1] as part of Population Health Improvement UK (PHI- UK), a national research network that seeks to transform health and reduce inequalities through change at the population level. S.L.H, R.R, J.O and H.W are supported by the Wellcome Trust [308556/Z/23/Z], S.L.H. currently receives funding from Impact on Urban Health part of Guy’s & St Thomas’ Foundation [EIC210605 and EIC221208], the Swedish Research Council [2023-05959] and the Wellcome Trust [28117/Z/23/Z & 223486/Z/21/Z]. The funders had no involvement in study design, data collection, analysis, interpretation or the decision to submit for publication. The views expressed are those of the author(s) and not necessarily those of the funders.

ZC prepared and submitted relevant materials for ethical approval. ZC recruited participants and drafted the manuscript alongside RR and HW with input from SH, LV, JO and LE. HW provided technical support and added necessary revisions to the manuscript in addition to acting as the corresponding author. All authors read and approved the final manuscript.

**Conflicts of Interest** The authors have no conflicts of interest to declare.

### Competing Interest Statement

The authors have declared no competing interest.

### Funding Statement

This paper represents independent research funded by the Wellcome Trust [203380/Z/16/Z] and the Economic and Social Research Council [ES/V009931/1]. J.O. is part-funded by the NIHR Biomedical Research Centre [BRC121520018] at South London and Maudsley NHS Foundation Trust. S.L.H. and R.R. are supported by the Economic and Social Research Council Centre for Society and Mental Health at King's College London [ES/S012567/1] and by UK Research and Innovation [MR/Y030788/1] as part of Population Health Improvement UK (PHI-UK), a national research network that seeks to transform health and reduce inequalities through change at the population level. S.L.H, R.R, J.O and H.W are supported by the Wellcome Trust [308556/Z/23/Z], S.L.H. currently receives funding from Impact on Urban Health part of Guy's & St Thomas' Foundation [EIC210605 and EIC221208], the Swedish Research Council [2023-05959] and the Wellcome Trust [28117/Z/23/Z & 223486/Z/21/Z]. The funders had no involvement in study design, data collection, analysis, interpretation or the decision to submit for publication. The views expressed are those of the author(s) and not necessarily those of the funders.

### Author Declarations

The Kings College London Research Ethics Committee for Psychiatry, Nursing and Midwifery and the NHS Health Research Authority gave ethical approval for this work (KCL Ref: HR-17/18-4629; HRA Ref: 18/HRA/0368)

## References

1. Schmitt, M.T., et al., The consequences of perceived discrimination for psychological well-being: a meta-analytic review. Psychological Bulletin, 2014. 140(4): p. 921.

2. Truong, M., Y. Paradies, and N. Priest, Interventions to improve cultural competency in healthcare: a systematic review of reviews. BMC Health Services Research, 2014. 14(1): p. 1.

3. Mays, V.M., et al., Perceived discrimination in healthcare and mental health/substance abuse treatment among blacks, latinos, and whites. Medical Care, 2017. 55(2): p. 173.

4. Van Ryn, M. and J. Burke, The effect of patient race and socio-economic status on physicians’ perceptions of patients. Social Science & Medicine, 2000. 50(6): p. 813–828.

5. Hugo, M., Mental health professionals’ attitudes towards people who have experienced a mental health disorder. Journal of Psychiatric and Mental Health Nursing, 2001. 8(5): p. 419–425.

6. Wigton, R.S. and W.C. McGaghie, The effect of obesity on medical students’ approach to patients with abdominal pain. Journal of General Internal Medicine, 2001. 16(4): p. 262–265.

7. Lateef, F., Simulation-based learning: Just like the real thing. Journal of Emergencies, Trauma and Shock, 2010. 3(4), pp. 348–352.

8. Cook, D.A., Hatala, R., Brydges, R., Zendejas, B., Szostek, J.H., Wang, A.T., Erwin, P.J. & Hamstra, S.J., Technology-enhanced simulation for health professions education: a systematic review and meta-analysis. JAMA, 2012. 306(9), pp.978–988.

9. Parsons, T.D., Neuropsychological assessment using virtual environments: enhanced assessment technology for improved ecological validity. In Advanced Computational Intelligence Paradigms in Healthcare 6. Virtual Reality in Psychotherapy, Rehabilitation, and Assessment, Springer, 2011, pp.271–289.

10. Dankbaar, M.E.W., Alsma, J., Jansen, E.E.H., van Merrienboer, J.J.G., van der Velden, T. & Schuit, S.C.E., An experimental study on the effects of a simulation game on students’ clinical cognitive skills and motivation. Advances in Health Sciences Education, 2016. 21(3), pp.505–521.

11. Kyaw, B.M., Saxena, N., Posadzki, P., Vseteckova, J., Nikolaou, C.K., George, P.P. & Car, J., Virtual reality for health professions education: systematic review and meta-analysis by the Digital Health Education Collaboration. Journal of Medical Internet Research, 2019. 21.

12. Slater M., Place illusion and plausibility can lead to realistic behaviour in immersive virtual environments. Philosophical Transactions of the Royal Society B, 2009. 364, 3549–3557.

13. Stone, J. & Moskowitz, G.B., Non-conscious bias in medical decision making: what can be done to reduce it? Medical Education, 2011. 45(8), pp.768–776.

14. Zestcott, C.A., Blair, I.V. & Stone, J., Examining the presence, consequences, and reduction of implicit bias in health care: a narrative review. Group Processes & Intergroup Relations, 2016. 19(4), pp.528–542.

15. Fung, K., Silver, D., Zhou, K., Sklar, M. & O’Sullivan, P., Use of virtual reality in the assessment of bias and empathy in medical students. Academic Psychiatry, 2019. 43, pp.514– 518.

16. Bohil, C.J., Alicea, B., Biocca, F.A., Virtual reality in neuroscience research and therapy. Nature Reviews Neuroscience, 2011. 12, 752–762.

17. Fung, K., Martin, L., George, R. & Arya, A., Exploring virtual reality in medical education: a review of the current landscape. Medical Education, 2019. 53(11), pp.1179–1189.

18. Peck, T. C., Seinfeld, S., Aglioti, S. M., & Slater, M. (2013). Putting yourself in the skin of a black avatar reduces implicit racial bias. Consciousness and cognition, 22(3), 779–787.

19. van Gelder, J.-L., M. Otte, and E.C. Luciano, Using virtual reality in criminological research. Crime Science, 2014. 3(1): p. 1.

20. Das-Munshi, J., D. Bhugra, and M.J. Crawford, Ethnic minority inequalities in access to treatments for schizophrenia and schizoaffective disorders: findings from a nationally representative cross-sectional study. BMC Medicine, 2018. 16(1): p. 1–10.

21. Tamayo-Sarver, J.H., et al., Racial and ethnic disparities in emergency department analgesic prescription. American Journal of Public Health, 2003. 93(12): p. 2067–2073.

22. Rhead, R.D., et al., Impact of workplace discrimination and harassment among National Health Service staff working in London trusts: results from the TIDES study. BJPsych Open, 2021. 7(1).

23. Riches, S., et al., Using virtual reality to assess associations between paranoid ideation and components of social performance: a pilot validation study. Cyberpsychology, Behavior, and Social Networking, 2019. 22(1), p.51–59.

24. Riches, S., et al., Factors affecting sense of presence in a virtual reality social environment: a qualitative study. Cyberpsychology, Behavior, and Social Networking, 2019. 22(4), p.288– 292.

25. Kourtesis, P., et al., Guidelines for the Development of Immersive Virtual Reality Software for Cognitive Neuroscience and Neuropsychology. 2021.

26. Radianti, J., Majchrzak, T.A., Fromm, J. & Wohlgenannt, I., A systematic review of immersive virtual reality applications for higher education: Design elements, lessons learned, and research agenda. Computers & Education, 2020. 147, 103778.

27. Slater M. & Sanchez-Vives M. V., Enhancing Our Lives with Immersive Virtual Reality. Frontiers in Robotics and AI, 2016. 3, 74.

28. Silverman, J. and P. Kinnersley, Calling time on the 10-minute consultation. British Journal of General Practice, 2012, p.118–119.

29. Rebenitsch, L. & Owen, C., Review on cybersickness in applications and visual displays. Virtual Reality, 2016.

30. StataCorp, Stata Statistical Software: Release 17. 2021, StataCorp LLC, College Station, TX.

31. Sevdalis, N., et al., Diagnostic error in a national incident reporting system in the UK. Journal of Evaluation in Clinical Practice, 2010. 16(6), p.1276–1281.

32. Panesar, S.S., et al., How safe is primary care? A systematic review. BMJ Quality & Safety, 2016. 25(7), p.544–553.

33. Chauhan, A., et al., The safety of health care for ethnic minority patients: a systematic review. International Journal for Equity in Health, 2020. 19, p.1–25.

34. Green, A.R., et al., Implicit bias among physicians and its prediction of thrombolysis decisions for black and white patients. Journal of General Internal Medicine, 2007. 22: p.1231–1238.

35. Penner, L.A., et al., The effects of oncologist implicit racial bias in racially discordant oncology interactions. Journal of Clinical Oncology, 2016. 34(24), p.2874.

